# Within-family attenuation of polygenic risk score accuracy: Investigating the effects of principal component analysis, LD score regression, and mixed model association in the UK Biobank

**DOI:** 10.1101/2025.08.28.25334633

**Authors:** Ciaran Michael Kelly, Onyedika Onuorah, Edmund Gilbert

**Author notes:** Corresponding author: (CMK).

## Abstract

A central challenge in the field of polygenic risk prediction has been measuring and controlling for confounding effects mediated through population stratification. Traditionally, such control has been attempted through the inclusion of the top principal components (PCs) of variation and the use of linear mixed models in genome-wide association studies (GWAS). Reductions in test-statistic inflation, commonly assessed using the genomic inflation factor (*λ*) and the linkage disequilibrium score regression (LDSC) intercept, as well as the preservation of polygenic risk score (PRS) predictive performance in within-family settings, are often taken as evidence that such confounding has been adequately controlled.

In this study, we examine the relationship between the number of PCs included during GWAS/PRS model development and the observed attenuation in performance when moving from a population-level setting to a cohort composed of discordant sibling pairs (within-family attenuation). The design enables the detection of confounding attributable to the environment and genetic background effects correlated with population structure. Analyses were conducted in the self-described White subset of UK Biobank (UKB) for coronary artery disease, type 2 diabetes, breast cancer, and prostate cancer. Educational attainment was included as a comparison trait, as it is known to exhibit substantial within-family attenuation.

We find that increasing the number of included PCs does not consistently reduce within-family attenuation among the traits examined. Moreover, reductions in attenuation do not closely track decreases in *λ* or the LDSC intercept, and the use of mixed model-based approaches provides little additional benefit for prediction or attenuation. These patterns persist even in settings where GWAS test-statistic inflation has been substantially reduced.

Our results suggest that the confounding in PRS targeted by PC-adjustment and linear mixed models is either minimal in the self-described White subset of the UKB or persists in ways that are not adequately captured by current population structure adjustment methods. Taken together, our findings suggest limitations as to how much standard population structure adjustment methods, or reductions in test statistic inflation, are currently improving the causal validity of PRS built using the UK Biobank.

## Introduction

Genome-wide association studies (GWAS) have been performed on many human complex traits and diseases to associate single-nucleotide polymorphisms (SNPs) with phenotypes of interest [1]. The summary statistics of a GWAS include an effect size and P-value for each examined SNP. These summary statistics can be used to compute a polygenic risk score (PRS) in an external cohort by weighting each individual’s allele dosages by the estimated SNP effects and summing across variants [2]. The potential ability of these PRSs to explain a large part of population-level variation of human disorders offers an important opportunity for personalized medicine. A polygenic score developed for coronary artery disease (CAD) demonstrates this potential. It identified individuals in the tail end of the risk score distribution that are at a three-fold higher risk of CAD than the general population, where this level of increased risk is comparable to that observed in individuals with monogenic forms of CAD [3]. Thus, those individuals at high risk may be offered prophylactic treatment options, such as statins in the case of CAD, or preventative lifestyle interventions for other conditions such as Type 2 Diabetes (T2D) [4, 5]. PRS have also shown to be substantial additional modifiers of disease risk or severity for several monogenic diseases such as diabetes or kidney disease [6, 7]. Even in the cases where individual-level prediction is not accurate enough to justify prophylactic treatment, PRS could be an efficient and cost-effective population-level tool for choosing earlier ages of screening for disease, such as in the case of breast and prostate cancer [8, 9]. Therefore, there is much interest in the use of polygenic scores as a central tool in precision medicine.

One of the major challenges when associating a SNP with any complex phenotype is in determining the source of the association. An observed association between a SNP and a phenotype can be attributed to: direct genetic effects; indirect genetic effects (including parental and sibling effects); and confounding effects [10]. Direct genetic effects are causal relationships between that variant and the trait of interest, for example, a SNP’s direct impact on gene expression that raises or lowers disease risk. Indirect genetic effects are the effects of others’ genotypes on an individual’s phenotype, operating through the environment they create or modify. For example, a parental allele that increases the parent’s own smoking behaviour can raise the child’s exposure to pro-smoking norms, increasing the child’s propensity to smoke. This reflects an indirect genetic effect via the parental environment, beyond any direct effect of the child’s own genotype. Confounding effects are spurious SNP–phenotype associations arising not from causal pathways of the SNP on the phenotype, but instead because the genotype is correlated with other variables that influence the phenotype. Confounding effects can be attributed to three primary sources: environment; genetic background; and assortative mating [10].

Environmental and genetic background confounding effects are often mediated through population structure in the data [11, 12]. Ideally, the SNP effect size estimates included in a PRS would be reflective of causal genetic effects, but if this population structure is not properly accounted for, confounding may remain. Therefore, developing techniques to measure and reduce confounding has been a central task in the genomic prediction literature, with debate still remaining on how best to detect and minimize these unwanted effects [13–16].

Perhaps the most commonly employed method for dealing with population structure has been the inclusion of principal components (PCs) of variation as covariates in GWAS. Principal component analysis (PCA) is a dimensionality reduction technique that generates orthogonal (i.e. linearly uncorrelated) components, each representing a distinct axis of genetic variation. In PCA, each successive PC explains progressively less variance than the previous.

The first PCs of a genomic SNP dataset often capture broad ancestry differences in the population, which can be used to control for confounding arising from population structure [13, 14, 17]. In the context of the UK Biobank, prior studies suggest that the first 16 PCs are sufficient for this purpose. These PCs have been used for several GWAS and PRS conducted on this dataset in the wider literature [18–20]. Although PC-based methods may be sufficient to control for genetic background confounding and environmental confounding correlated with ancestry, it is not thought to contribute to removing associations driven by indirect genetic effects or assortative mating [21, 22].

The addition of mixed model approaches to GWAS has now become common [23, 24]. These methods employ a linear mixed model (LMM) and are commonly referred to as mixed-linear models of association (MLMAs). In this procedure, a genetic relatedness matrix (GRM) is constructed and is used to model the covariance structure of the random effect in the LMM. MLMA approaches are thought to better control for population stratification and cryptic relatedness than PC-adjustment alone. The MLMA approach may also provide some control for confounding due to assortative mating [22]. Extensions appropriate for binary phenotypes through the use of generalized linear mixed models (GLMMs) have also been developed [25].

Determining the success of methods that attempt to control for confounding in GWAS is challenging. The genomic inflation factor (*λ*) is a measure of global P-value inflation in the summary statistics outputted from a GWAS [26–29]. When *λ* is significantly above one (i.e. P-values are systematically lower than the expectation under the null) this suggests that population stratification may not have been successfully corrected for, or that the trait is highly polygenic [30]. The intercept and ratio produced by linkage disequilibrium score regression (LDSC) is thought to distinguish confounding signal from that of true polygenicity [30, 31]. The LDSC intercept provides an estimate of the absolute component of test statistic inflation that is consistent with confounding (with values close to 1 indicating minimal confounding), and the ratio value expresses this same quantity relative to the total inflation in the data, i.e. the proportion of inflation attributable to confounding rather than polygenic signal (with values ideally close to zero). Despite many such control and measurement techniques, some authors argue that the current methods may not yet be sufficient to truly correct for all the environmental and genetic background confounding present in GWAS effect size estimates [10, 16, 32]. As discussed above, much debate has surrounded the causal nature of the genetic effects estimated from GWAS and included in PRS models [11, 12, 33, 34]. This has been compounded by the fact that PRS models are often constructed with SNPs which fall well below the canonical genome-wide significance threshold, plausibly increasing the risk that genetic background and environmental confounding effects will be exploited during prediction [2, 35].

One method that has been proposed as critical to validating the causal nature of a given polygenic risk score has been the within-family study design. PRS trained on population data are expected to show reduced discrimination in within-family tests i.e. their predictive ability has been attenuated. This attenuation occurs for two principal reasons: (i) family members share much of their SNP profile, which reduces PRS variation within families; and (ii) the within-family design removes signal due to population stratification, assortative mating, and most indirect genetic effects (with the exception of some sibling indirect effects) [36–38]. The degree to which a PRS discriminates between relatives (often siblings), as compared to non-relatives in a population sample, is therefore informative about the proportion of the given PRS reflecting direct genetic effects [16, 39]. It must be stated that even if perfect control of genetic and environmental confounding is achieved when building a PRS, residual attenuation is still expected from assortative mating and indirect effects, and from the aforementioned reduced genetic variation among family members.

As predictive discrimination between siblings can be ascribed to the influence of causal genetic effects, measuring changes in predictive attenuation during population stratification control is a valuable approach. PCA- and LMM-based methods target the genetic background and environmental component of confounding only. Therefore, if such confounding is a significant source of population-level performance, applying such adjustments should reduce the predictive attenuation observed when moving to a within-family setting. If attenuation does not reduce after adjustment, it can be inferred that this form of confounding was minimally important for the PRS, or that it is persisting in forms not well-captured by standard PCs and LMMs.

The approach taken in this paper follows from the work of Lello et al. who have previously developed polygenic risk scores from the UK Biobank and validated them in a within-sibling cohort [38]. The authors found that a significant amount of polygenic predictive power remains when moving from a population-level cohort to a within-family setting. However, the authors did not make use of principal components or mixed models in the development of their PRS on the basis that their top PCs do not explain a substantial amount of phenotypic variation in the context of human height [40].

Extending this previous work, our study systematically evaluates how including principal components and using mixed models during GWAS and PRS construction affects within-sibling predictive attenuation across multiple complex traits. This design isolates attenuation attributable to population structure and ancestry-correlated confounding. Reductions in predictive attenuation observed with PC- and LMM-based adjustments likely reflect improved control of these factors, whereas remaining attenuation could stem from residual background or environmental confounding not captured by these methods, as well as from sources such as shared genetics, assortative mating, and indirect genetic effects. As the remaining association between a PRS and a phenotype in a within-family setting can be attributed to direct genetic effects (with at most a small contribution from sibling indirect effects), the overall aim was to assess whether the commonly used methods for controlling for stratification appear to meaningfully impact the causal validity of PRS built using the UK Biobank.

## Materials and methods

### Study approach

We performed GWAS in the UK Biobank and constructed PRS to compare predictive performance between two types of disease-discordant pairs: unrelated case–control pairs and sibling case–control pairs. As principal components must be included as covariates at the GWAS stage, separate GWAS were performed for each level of PC-adjustment, generating distinct sets of summary statistics for downstream PRS construction. We assessed attenuation in classification accuracy when moving from population-level prediction to within-family prediction, and examined how varying the population structure adjustment strategy and SNP-set sizes influenced this attenuation.

### Phenotype selection

Phenotype definitions followed those used in Lello et. al. (2020) with the exception of educational attainment which was converted to a pseudo-binary phenotype in order to facilitate comparisons in PRS performance and attenuation [38]. Dichotomizing educational attainment in this manner sacrifices information and interpretability and we employ it only to benchmark comparisons with the binary disease outcomes in terms of confounding. The precise encoding of each phenotype can be found in Supplementary note 1.

### Population selection and genotype quality control

As per Lello et al., all individuals with ethnic background codes 1, 1001, 1002, or 1003 (i.e. self-reported “White” individuals) were included in the GWAS and PRS analyses [38]. The following quality control (QC) measures were implemented using PLINK1.9: minor allele frequency (MAF) threshold of 0.1%, SNP missingness filter of 3%, and individual missingness filter of 3%. The extended human leukocyte antigen (HLA) region and sex chromosomes were also excluded [42].

### Discordant pair extraction

#### Siblings

Siblings were defined as those pairs which have an IBS0 *>*0.0012 and kinship coefficient *>*0.176. For each trait, all sibling pairs in which one individual was a case and the other a control were identified. Pairs were kept single-sex for prostate and breast cancer. Trios and higher-order relationships were not included in the analysis i.e. only the first pair were kept when more than one sibling pair was identified within a single family unit.

#### Non-siblings

After the creation of the discordant sibling test sets as described above, a KING relatedness threshold of *≥* 0.0884 was implemented to exclude closely related individuals from the post-QC dataset. Random case-control pairs were selected from the unrelated cohort, with sample sizes matched to the corresponding sibling test sets. For prostate and breast cancer, pairs were restricted to individuals of the same sex.

To match the age difference distribution observed in the sibling pairs, candidate non-sibling pairs were first stratified into equivalent age difference bins. Proportional sampling was then performed within these bins to replicate the sibling age difference distribution. The resulting set was trimmed or supplemented to match the exact number of sibling pairs, thus ensuring balanced age distributions and sample sizes between groups.

### Principal component analysis

Principal components were taken from UK Biobank data field 22009 and individuals with missing PC information were excluded (n=14,227). The first 16 principal components have previously been stated to be sufficient to capture population structure in the UK Biobank [41].

### Association methods

#### Standard GWAS (GLM)

PLINK2’s --glm function was used to perform basic GWAS across the phenotypes [43]. The discordant test sets made up of sibling and non-sibling pairs were excluded from the training GWAS so as not to bias results. To investigate the effect of including varying numbers of principal components of genetic variation, the top *M* PCs were included as covariates during association. Separate GWAS were therefore performed for each value of *M* , generating distinct summary statistics for PRS construction.

#### Mixed model GWAS

GCTA software was used to test for the added effect of performing a mixed model-based approach to controlling for population stratification [44]. The post-QC SNPs were pruned using PLINK1.9 for construction of the GRMs with the following parameters: window-size of 100kb, step-size of 5bp and squared correlation threshold of 0.5 [42]. The HLA region was excluded during pruning. GRMs were constructed using GCTA version 1.92.3 on the resultant set of 111,226 SNPs. A sparsity threshold of 0.05 was then applied to the GRM. A GCTA 1.94.4 generalized linear mixed model association analysis (fastGWA-GLMM) was then performed on each trait with the full set of 16 principal components of variation included as covariates [25].

#### Clumping

SNPs were clumped using PLINK1.9 [42]. The LD threshold for clumping was set to 0.1, the distance threshold to 500kb, and the index SNP significance threshold to 0.5.

### LD-score regression

LD-Score Regression (LDSR) was implemented to distinguish test statistic inflation due to polygenicity from that of confounding [30, 31]. The regression intercept was estimated using the ldsr software v.1.01. Reference linkage disequilibrium (LD) scores and weights specific to those of European ancestries from the 1000 Genomes Project reference panel were used for this analysis [45]. The HLA region was excluded from the LD score reference files.

### Polygenic score construction

Polygenic scores were constructed for the discordant test set made up of sibling and non-sibling pairs with the top *N* clumped SNPs and using the --score function of PLINK2 [43]. Raw scores were regressed on the top *M* principal components of genetic variation. The resulting residuals were used as the predictor variable for the phenotype of interest.

### Classification

The outcome variable for classification of discordant pairs was defined as the proportion of pairs in which the positive-class individual had the higher polygenic score (with ties excluded) [38].

This metric is conceptually similar to the area under the receiver operating characteristic curve (AUC) which represents the probability of correctly assigning a higher score to the case in a randomly selected discordant pair [46]. A major difference between the AUC and the classification metric used here is that the AUC represents an overall probability from all possible randomly selected discordant pairs in the population. The metric used here is based on a specific pre-defined and non-random set of pairs composed of the siblings and non-siblings.

Attenuation in performance was defined as the proportional reduction in above-chance classification accuracy when moving from unrelated pairs to siblings. An attenuation of 0% indicates no loss i.e. the PRS performed equally well in siblings and non-siblings. 100% attenuation corresponds to a complete loss of predictive power in the within-family setting. Negative attenuation values can arise in the rarer instance where there is a greater predictive power in the sibling pairs as compared to the non-sibling pairs.

### Statistical analysis

A generalized linear mixed model (GLMM) was fit to investigate the overall effects of pair type, adjustment method, phenotype, and SNP-set size on classification accuracy. Three adjustment methods were compared: 0-PC, 16-PC, and GLMM+16-PC. An interaction term between method and pair type was included to assess whether the performance of the methods differed between sibling and non-sibling pairs. To account for repeated use of the same case-control pairs, a random intercept was included for each pair. The lme4 package was used to implement this GLMM using the BOBYQA optimizer [47, 48].

## Results

### Datasets and traits analysed

Analyses were conducted using the 2018 UK Biobank genotype release in the self-reported White ancestry subset. Principal component analysis (PCA) plots are shown in Supplementary Figures S1 and S2. We examined four disease traits for which polygenic risk scores have shown potential clinical utility [49, 50]. For the purposes of comparison, we examined the effects of controlling for population stratification on a binarized version of the educational attainment phenotype which has been well-established as having issues with confounding and within-family attenuation [22, 37, 38]. The relative importance of contributing factors to this attenuation is not currently settled, with some suggestion that population stratification may play a major role [51, 52]. Other authors have also found a large role for indirect genetic effects in this trait [36, 53].

Summary statistics from each GWAS were used to build PRS models used to classify between discordant pairs (consisting of one case and one control in each pair). Two test sets were evaluated, one consisting of sibling pairs and the other of population-level unrelated pairs (non-siblings). To account for the effects of different levels of polygenicity, three SNP-set sizes were examined during PRS construction (1K, 10K, and 100K). The total sample size of each phenotype, and the discordant sibling and non-sibling test sets can be seen in Table 1 and 2 respectively.

**Table 1.**
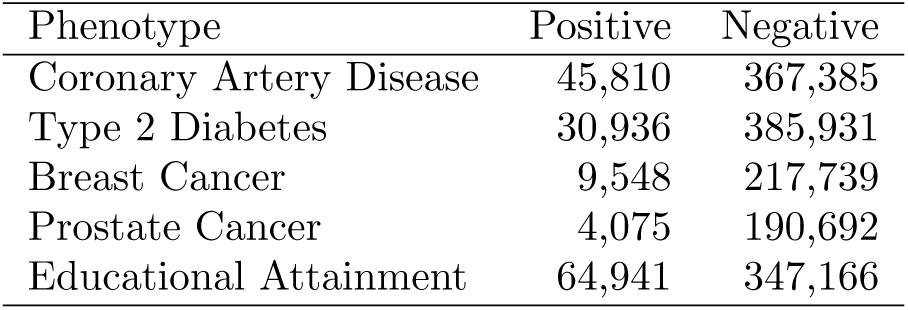
Sample sizes in training GWAS for positive and negative phenotype classes in the UK Biobank.

The primary outcome was the attenuation in predictive performance observed when moving from population-level to within-family prediction.

### Baseline PRS accuracy and attenuation

To establish baseline performance metrics and quantify the degree of confounding present prior to population structure adjustment, we first constructed PRSs using standard GLM-based GWAS without controlling for population stratification (i.e. no PCs were included as covariates). The predictive accuracy of these PRSs in both the population-level and within-family settings is presented in Table 3. Overall, predictive accuracy was modest across all traits, with no PRS exceeding 65% classification accuracy in the population-level setting. As expected, we observed reduced accuracy among discordant sibling pairs compared to unrelated individuals across most trait-SNP set combinations, reflecting within-family attenuation. Educational attainment exhibited the largest attenuation, with the majority of classification accuracy lost when transitioning from the population-level to within-family setting, consistent with prior literature documenting substantial confounding in this trait [22, 37, 38].

**Table 2.** The number of individuals in discordant sibling test sets from the UK Biobank. Additional test sets made up of an equal number of unrelated population-level pairs were generated.

| Phenotype | Number of Siblings |
| --- | --- |
| Coronary Artery Disease | 7,512 |
| Type 2 Diabetes | 5,024 |
| Breast Cancer | 1,196 |
| Prostate Cancer | 294 |
| Educational Attainment | 8,280 |

**Table 3.** Baseline classification accuracy and attenuation for standard GWAS-based PRS performance on sibling and population-level (non-sibling) discordant pairs in the UK Biobank. No principal components were included as covariates in either the GWAS or PRS.

| Phenotype | SNP-set size | Classification accuracy (%) |  | Attenuation (%) |
| --- | --- | --- | --- | --- |
|  |  | *Non-sibling pairs* | *Sibling pairs* |  |
| Coronary Artery Disease | **1K** | **57.29** | **53.78** | **48.1** |
|  | 10K | 56.58 | 55.40 | 17.9 |
|  | 100K | 55.99 | 55.01 | 16.4 |
| Type 2 Diabetes | 1K | 62.46 | 59.28 | 25.5 |
|  | **10K** | **63.54** | **59.00** | **33.5** |
|  | 100K | 63.54 | 58.52 | 37.1 |
| Breast Cancer | **1K** | **58.03** | **54.85** | **39.6** |
|  | 10K | 56.52 | 53.68 | 43.6 |
|  | 100K | 55.02 | 54.01 | 20.1 |
| Prostate Cancer | **1K** | **58.50** | **60.54** | **-24.0** |
|  | 10K | 57.82 | 55.78 | 26.1 |
|  | 100K | 57.14 | 54.42 | 38.1 |
| Educational Attainment | 1K | 58.33 | 51.93 | 76.8 |
|  | **10K** | **59.76** | **53.48** | **64.3** |
|  | 100K | 59.69 | 53.14 | 67.6 |

Among the disease traits, attenuation was generally significant but more moderate in magnitude. A notable exception to this was that the 1,000 SNP prostate cancer model showed negative attenuation, with the PRS demonstrating higher predictive accuracy in the sibling set than among unrelated individuals. This pattern was not observed for other SNP set sizes and may be reflective of stochastic variation.

We did not observe a consistent relationship between SNP set size and either predictive accuracy or attenuation magnitude. Specifically, larger SNP sets did not consistently yield improved prediction in the population-level setting, nor did they exhibit increased attenuation as might be expected if the inclusion of SNPs with lower p-values was more susceptible to capturing confounding effects. Based on performance in the population-level test set, we selected the best-performing SNP set size for each

trait to carry forward in subsequent analyses examining the impact of PC adjustment (bolded in Table 3).

### Changes in the LDSC intercept with increasing PC inclusion

To evaluate whether standard population structure adjustment methods effectively reduce confounding as traditionally measured, we examined the effect of including an increasing number of PCs on the LDSC intercept and ratio across all traits. The LDSC intercept results are presented in Figure 1, with corresponding LDSC ratio trends shown in Supplementary Figure S4. The intercept decreased for most traits as the number of PCs included in the training GWAS increased, though the magnitude and pattern of reduction varied substantially across phenotypes.

**Fig 1.**
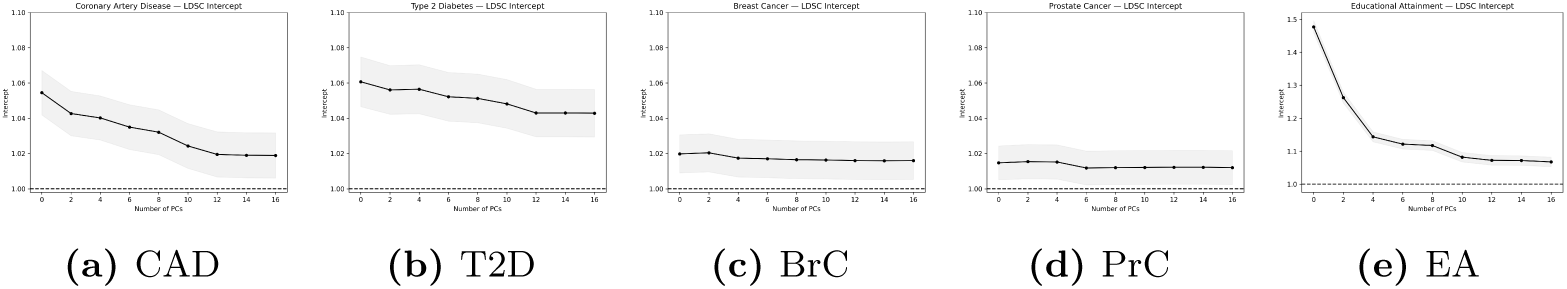
Changes in the LD score regression (LDSC) intercept with increasing inclusion of principal components (PCs) across phenotypes: coronary artery disease (CAD), type 2 diabetes (T2D), breast cancer (BrC), prostate cancer (PrC), and binarized educational attainment (EA). LDSC summary statistics were derived from GLM-based GWAS results. Shaded regions represent standard errors. *Note:* The y-axis limit is higher for educational attainment due to greater overall inflation.

Consistent with the baseline attenuation results, educational attainment exhibited the strongest response to PC-adjustment. Both the intercept and ratio were substantially elevated at baseline (no PCs included), indicating considerable confounding. While subsequent inclusion of PCs produced marked reductions in both metrics, residual confounding likely remained even when all sixteen principal components were included as covariates [54]. Coronary artery disease and type 2 diabetes demonstrated moderate baseline evidence of confounding, with intercepts modestly elevated above 1.0. Both traits showed downward trends in the intercept as more PCs were included, though the absolute magnitude of change was not extreme. Notably, after PC correction with the full complement of 16 PCs, both traits achieved LDSC ratios below 1.1, falling under a rule-of-thumb threshold for minor confounding [55]. Breast and prostate cancer exhibited low absolute levels of confounding at baseline. The LDSC ratios for these traits were more moderate but associated with large standard errors, possibly reflecting sample size limitations for these two phenotypes. Finally, similar patterns as these trends were observed for the genomic inflation factor *λ* across all traits (Supplementary Figure S3).

### PRS accuracy across levels of PC inclusion

To assess whether reductions in test statistic inflation translate to changes in predictive performance, we constructed PRSs for each phenotype across all levels of PC inclusion, with PCs included as covariates in both the training GWAS and during PRS prediction.

Results for the best-performing SNP set size for each trait are presented in Figure 2. Among unrelated individuals, we observed no consistent monotonic relationship between the number of included PCs and predictive accuracy, though modest variation across PC levels was evident. Among discordant sibling pairs, there was a slight overall trend toward improved performance with increasing PC inclusion, though this pattern was not universal across all traits.

**Fig 2.**
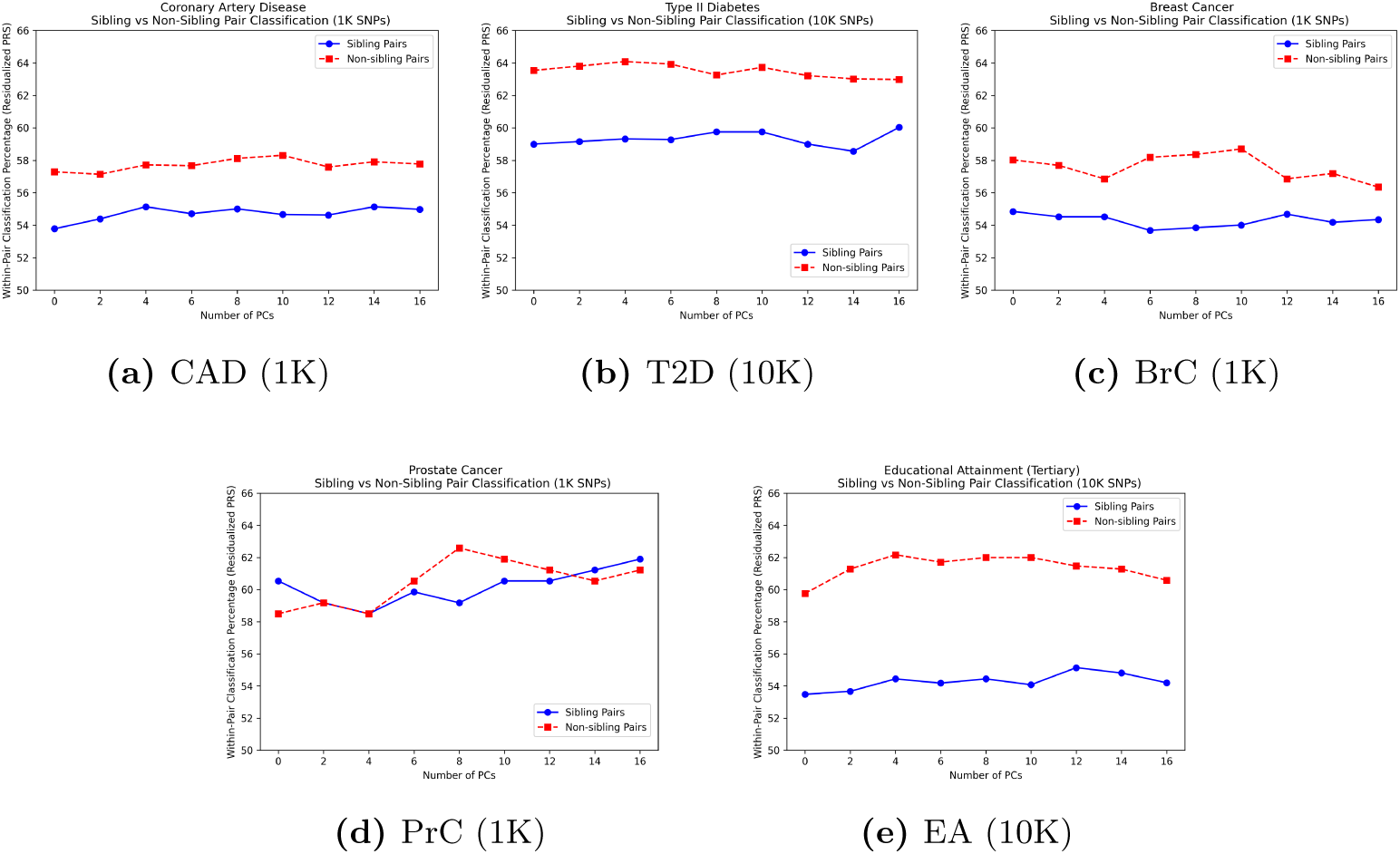
PRS classification accuracy across levels of PC inclusion for sibling and non-sibling discordant pairs, shown for the best-performing SNP-set size per phenotype.

When examining the full range of SNP set sizes, the absence of a consistent trend in either unrelated individuals or sibling pairs became even more apparent (Supplementary Figures S5–S9).

The principal components used in these analyses were computed across the entire UK Biobank cohort, including individuals of more diverse ancestry than those included in our ancestry-filtered GWAS. To assess whether this might diminish the effectiveness of PC-adjustment for our analysis cohort, we recomputed PCs using only individuals from the post-QC, self-described White subset. When these cohort-specific PCs were used in GWAS and PRS construction, they did not substantially alter predictive accuracy for any of the disease traits examined (Supplementary Tables S2 and S3).

Whilst overall predictive accuracy is informative, the primary outcome of interest for evaluating confounding is not absolute performance. PRS accuracy may reflect both causal genetic effects and confounding, whereas within-family attenuation directly quantifies the degree to which population-level associations fail to replicate in a setting that controls for both genetic background and environmental confounding.

### Predictive attenuation across levels of PC inclusion

We next examined whether PC-adjustment reduces within-family attenuation. Reductions in attenuation when transitioning from the population-level to within-family setting would indicate that PC adjustment successfully controls for environmental and genetic background confounding. The experimental design here allows us to isolate the specific effects of environmental and genetic background confounding that PC-adjustment is intended to target. Whilst some persistent attenuation is expected due to shared genetics between siblings and the other sources of confounding not addressed by PC-adjustment, reductions in attenuation with increasing PC inclusion can provide evidence that standard population structure adjustment methods are successfully mitigating the confounding they are specifically designed to control.

Attenuation results for each trait are presented in Figure 3. Across all traits, we did not observe a consistent decrease in attenuation as more PCs were included. A weak trend toward reduced attenuation may be present in some cases, although the relationship was neither monotonic nor uniform. Notably, attenuation sometimes increased or decreased in a non-systematic manner depending on the specific number of included PCs, suggesting that the optimal number of PCs for minimizing attenuation is not predictable in advance of within-family validation, or through observation of the LDSC intercept. This lack of consistent pattern was further confirmed when examining all SNP set sizes (Supplementary Figure S10).

**Fig 3.**
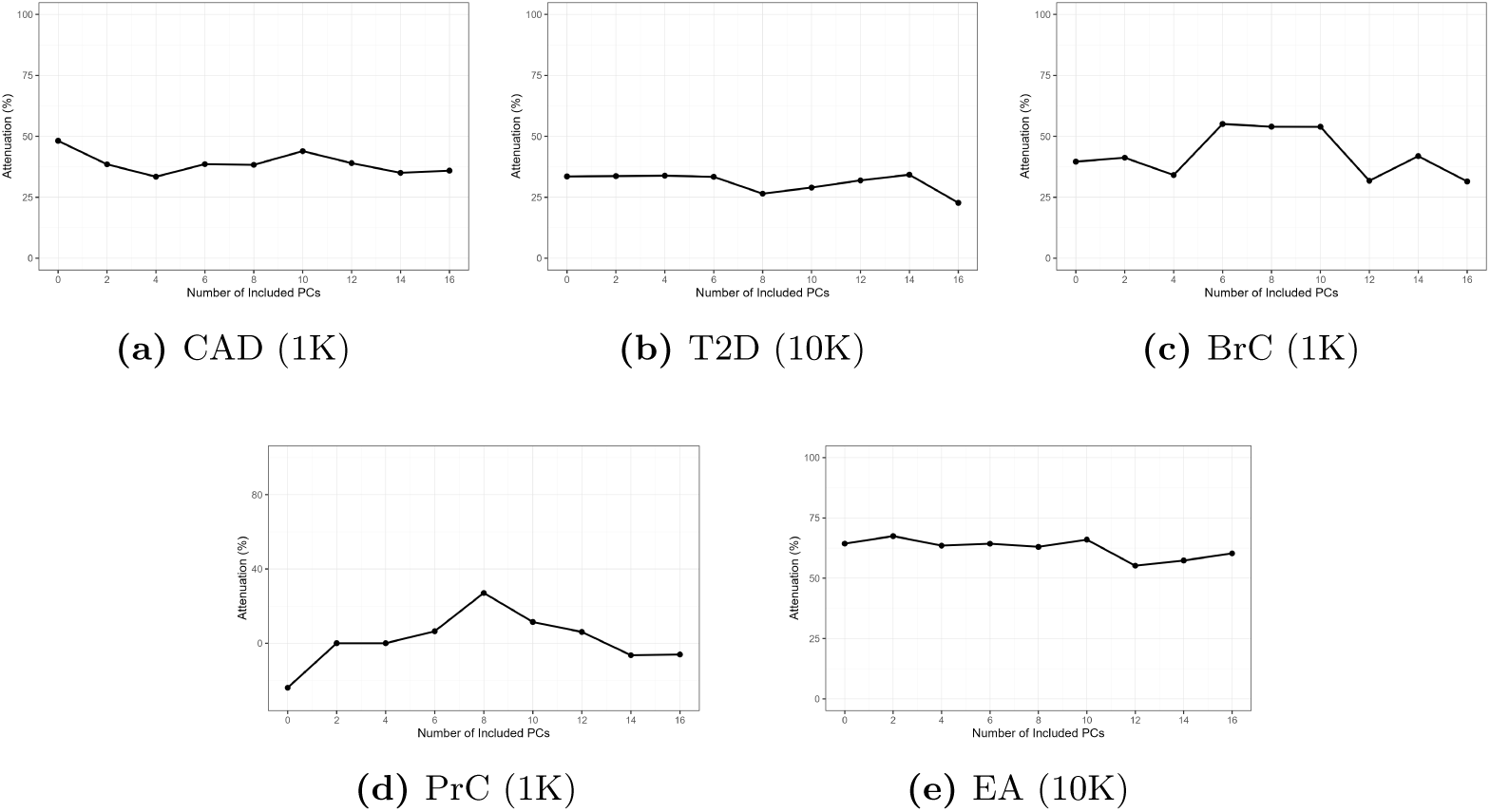
Attenuation in PRS predictive performance between sibling and non-sibling discordant pairs across levels of PC inclusion, shown for the best-performing SNP-set size per phenotype.

Educational attainment exhibited persistently high attenuation across all levels of PC inclusion, despite the substantial reduction in LDSC intercept documented above. Similarly, for coronary artery disease and type 2 diabetes, the decreases in LDSC intercept observed with increasing PC inclusion were not accompanied by corresponding reductions in within-family attenuation. These findings indicate that strong-to-moderate reductions in confounding as measured by the LDSC intercept do not translate to meaningful improvements in within-family predictive validity. This suggests some limitations in using test statistic inflation metrics as proxies for the causal validity of polygenic risk scores.

Our results are consistent with prior work suggesting that population structure (as captured by PCs) may contribute only modestly to PRS performance for some traits in the UK Biobank. For example, Lello et al. showed for height that regressing the phenotype on principal components yields low explanatory power, implying limited variance attributable to PC-defined structure [40]. Similarly, although several PCs were significantly associated with each phenotype in our data, we did not observe a clear relationship between the significant PCs and specific changes in predictive performance or within-family attenuation, despite the downward trends in overall inflation mediated by PC-adjustment (Supplementary Table S1).

### Additional effects of mixed model GWAS

To investigate whether GRM-based mixed models provide additional control over confounding beyond PC-adjustment alone, we performed GWAS using fastGWA-GLMM with the full set of 16 PCs included as covariates. We compared the resulting PRSs to those derived from standard GLM-based GWAS with either 0 PCs or 16 PCs.

Results for the best-performing SNP set size for each trait are presented in Table 4. As already shown in the previous findings, inclusion of the full set of PCs may have produced slight improvements in accuracy and reductions in attenuation relative to models with no PC adjustment. However, the addition of mixed model correction provided no apparent benefit beyond that achieved through PC inclusion alone. Within-family attenuation was nearly identical between the 16-PC GLM and GLMM+16-PC approaches across most traits examined.

**Table 4.** Classification accuracy and within-family attenuation for PRS derived from GWAS across population structure adjustment methods. Results are shown for sibling and non-sibling discordant pairs from the UK Biobank. For each phenotype, rows correspond to an unadjusted GLM-based GWAS (0-PC), a GLM-based GWAS including 16 PCs (16-PC), and a mixed model-based GWAS including 16 PCs (GLMM+16-PC).

| Phenotype | Method | Classification accuracy (%) |  | Attenuation (%) |
| --- | --- | --- | --- | --- |
|  |  | *Non-sibling pairs* | *Sibling pairs* |  |
| Coronary Artery Disease<br>(1K) | 0-PC | 57.29 | 53.78 | 48.1 |
|  | 16-PC | 57.77 | 54.98 | 35.9 |
|  | GLMM+16-PC | 57.72 | 54.98 | 35.5 |
| Type 2 Diabetes<br>(10K) | 0-PC | 63.54 | 59.00 | 33.5 |
|  | 16-PC | 62.98 | 60.03 | 22.7 |
|  | GLMM+16-PC | 63.02 | 59.83 | 24.5 |
| Breast Cancer<br>(1K) | 0-PC | 58.03 | 54.85 | 39.6 |
|  | 16-PC | 56.35 | 54.35 | 31.5 |
|  | GLMM+16-PC | 55.69 | 54.85 | 14.8 |
| Prostate Cancer<br>(1K) | 0-PC | 58.50 | 60.54 | -24.0 |
|  | 16-PC | 61.22 | 61.90 | -6.1 |
|  | GLMM+16-PC | 59.18 | 61.22 | -22.2 |
| Educational Attainment<br>(10K) | 0-PC | 59.76 | 53.48 | 64.3 |
|  | 16-PC | 60.58 | 54.20 | 60.3 |
|  | GLMM+16-PC | 60.72 | 54.18 | 61.0 |

This pattern was also largely consistent across all SNP set sizes (Tables 3, S5, and S6). Furthermore, the GLMM approach did not meaningfully reduce the genomic inflation factor *λ* beyond the level achieved with 16 PCs in a standard GLM framework (Supplementary Table S4). Taken together, these results suggest that in large and relatively homogeneous cohorts such as the UK Biobank, mixed model methods do not offer substantive improvements over PC-adjustment for controlling confounding effects attributable to population stratification.

To more formally assess the factors influencing classification accuracy, we fit a generalized linear mixed model (GLMM) with phenotype, SNP-set size, pair type (sibling vs. non-sibling), and adjustment method (0-PC, 16-PC, and GLMM+16-PC) as fixed effects. This analysis allowed the relative importance of each factor to be evaluated while accounting for repeated measurements across traits and models.

Phenotype emerged as the strongest predictor of classification accuracy (*χ*^2^ = 114, *p <* 1 *×* 10*^−^*^15^). Pair type was also strongly associated with accuracy (*χ*^2^ = 80.1, *p <* 1 *×* 10*^−^*^15^), reflecting the substantial reduction in predictive performance observed when moving from population-level to within-family comparisons. SNP-set size had a more modest but still statistically significant effect (*χ*^2^ = 21.6, *p* = 2 *×* 10*^−^*^5^).

In contrast, population structure adjustment method explained relatively little variation in predictive accuracy overall (*χ*^2^ = 9.6, *p* = 0.008), with both the 16-PC and GLMM+16-PC approaches yielding only slight improvements relative to the unadjusted model. Importantly, the interaction between adjustment method and pair type was not significant (*χ*^2^ = 0.008, *p* = 0.996), indicating that all three methods exhibited comparable levels of attenuation when transitioning from unrelated individuals to sibling pairs.

Together, these results suggest that more sophisticated population structure correction strategies do not substantially improve within-family transferability of PRS beyond standard PC adjustment in this dataset, and that reductions in population-level test-statistic inflation do not necessarily translate into gains in causal predictive validity.

## Discussion

Our study design enabled us to assess how effectively principal component adjustment and mixed models reduced within-family PRS attenuation attributable to genetic background and environmental confounding. Because predictive power in sibling-based analyses primarily reflects direct genetic effects, this framework allowed us to evaluate whether standard population stratification controls improve the causal validity of PRS models in the UK Biobank.

Several expectations regarding population stratification control in PRS development did not consistently hold across traits in this analysis. In particular, increasing the number of principal components included during GWAS and PRS construction did not reliably reduce within-family attenuation of predictive performance. Our formal statistical analysis confirmed that the PC-adjustment and mixed model approaches exhibited similar attenuation to the baseline approach when moving from unrelated to sibling pairs. Whilst a general reduction was observed in the genomic inflation factor *λ* and LDSC intercept with the inclusion of additional PCs, this did not consistently translate into improvements in the within-family prediction task. These findings suggest that reductions in test statistic inflation should not be interpreted as direct evidence of significantly improved causal validity of PRS models.

The use of linear mixed models for the generation of GWAS summary statistics also did not significantly reduce either *λ* or attenuation beyond that of PC-control alone. For several traits, breast and prostate cancer in particular, the effects of attempting stratification control appeared minimal regardless of the number of PCs included or the use of LMMs. This potentially reflects low baseline levels of confounding in these traits, or limited power to detect and minimize its effects. Even for educational attainment, which is thought to have substantial vulnerability to confounding, the success of PC or mixed model-based correction in fully accounting for stratification was questionable. Whilst the LDSC intercept and *λ* decreased with additional PCs as expected, a concomitant reduction in attenuation was not observed. Variation in SNP-set size also had little consistent effect on within-family attenuation or population-level performance across traits. This suggests that increased polygenicity does not necessarily exacerbate confounding effects or alter the effectiveness of stratification control in this context.

Even after substantial attempts at such stratification control, attenuation in prediction remained substantial in some traits. This may reflect some residual genetic background or environmental confounding and likely stems from other sources of attenuation, such as indirect parental effects and assortative mating. Our experimental design could not distinguish between these sources beyond the observed reductions associated with stratification control, which are typically attributed to genetic background and environmental confounding. Nonetheless, prior studies have demonstrated that indirect genetic effects and assortative mating can meaningfully contribute to attenuation in complex traits [22, 36, 56].

The relationship between test statistic inflation and within-family attenuation varied substantially across traits in ways that provide insight into the nature of the confounding present. For the binarized educational attainment phenotype, both the LDSC intercept and ratio were substantially elevated at baseline, indicating considerable confounding. This served as a useful benchmark to compare the disease phenotypes with. PC-adjustment produced marked reductions in both values, yet within-family attenuation remained persistently high. This dissociation suggests that the confounding captured by these metrics may not be especially relevant in terms of trait prediction.

For coronary artery disease and type 2 diabetes, baseline LDSC metrics were moderately elevated. PC-adjustment reduced both metrics, with ratios falling to acceptable levels (below 1.1) after inclusion of 16 PCs, though the overall magnitude of this reduction was not particularly large. Despite some improvements in test statistic inflation, a corresponding reduction in within-family attenuation was not observed. Attenuation for these traits did fluctuate depending on the specific number of PCs included, but no general downward trend was observed as more PCs were added. This lack of systematic relationship between PC inclusion and attenuation, despite measurable reductions in LDSC metrics, again suggests that the confounding captured and reduced by standard test statistic inflation measures may not correspond to the confounding affecting within-family prediction validity. Overall, however, LDSC statistic improvements were modest relative to educational attainment, limiting their potential impact on within-family attenuation.

Taken together, these findings suggest that current best practices, such as adjusting for a pre-defined fixed number of PCs, may not generalize well across phenotypes. Many GWAS and PRS models derived from the UK Biobank resource employ these or similar adjustment methods as default [18–20]. The present results, however, raise the question of whether these corrections are consistently useful in this cohort. It remains unclear whether the limited influence of stratification control on attenuation reflects genuinely minimal confounding within the UK Biobank or, conversely, confounding that persists in forms not adequately addressed by conventional methods. If the former is true, routine application of these corrections may add complexity without substantive benefit. If the latter is true, more sophisticated approaches will be required to capture subtle or residual population structure. Although we cannot determine definitively whether any remaining attenuation after adjustment is due to environmental or genetic background confounding, this possibility cannot be excluded and must still be considered.

Regardless of the underlying mechanism behind predictive power, be that genetically causal or not, polygenic risk scores that successfully discriminate higher-risk individuals at a population-level may still serve a useful function in clinical and public health contexts. Debate remains as to whether or not a lack of full causal interpretability necessarily precludes implementation of PRS as a screening or risk stratification tool [12, 33, 34, 57]. Although this study focused only on linear polygenic risk scores, the question of establishing causality may increase in importance if the field of genomic prediction shifts toward reliance on “black-box” machine-learning models such as neural networks [58, 59]. Non-linear models could exploit complex confounding patterns that cannot be adequately addressed through linear approaches such as PCA. For this reason, focus on the development of robust validation frameworks for assessing the causal interpretation of PRS may increase in the years ahead.

Several limitations should be considered when interpreting these findings. Firstly, the number of traits included in this analysis was small and the sample sizes of the test sets were relatively modest, especially for the two cancer phenotypes. The size of currently available within-family cohorts is limited, however, there do exist external independent datasets that could be used to potentially validate some of these findings [36].

Secondly, sibling indirect effects may marginally affect within-family PRS performance, meaning that predictive signal between siblings may not solely reflect strictly proximally biologically causal pathways. However, the present design specifically evaluated improvements in causal validity associated with population stratification control methods, which are not expected to mitigate sibling indirect effects. As such, these effects are unlikely to substantially influence the interpretation of our results.

Thirdly, the results shown here only apply to those of self-declared White ancestry and present in the UK Biobank. The nature of the recruitment processes and relatively small geographic catchment area might limit the extent to which environmental and genetic background confounding is an issue in the GWAS being conducted on this data. Analyses on other populations or more diverse cohorts could see much larger effects of stratification control, especially in those of admixed continental ancestry.

In conclusion, attempting to control for population stratification on polygenic risk scores derived from UK Biobank data warrants careful consideration. The relationship between test statistic inflation, principal component or mixed model adjustment, and the within-family attenuation is complex and not readily predictable. No single correction heuristic appears sufficient to maximize predictive performance and establish causality across all the traits examined. We therefore recommend incorporating direct within-family validation, when possible, to empirically inform the selection of population stratification control measures being used during model construction.

## Data Availability

This study was conducted using the UK Biobank resource under approved application
103770. Individual-level genotype and phenotype data from the UK Biobank are
available upon application to the UK Biobank. Data supporting the findings of this
study, including GWAS summary statistics and classification scripts, are available from the corresponding author upon request.

## Acknowledgments

This research has been conducted using the UK Biobank Resource under Application Number 103770. We thank all participants of the UK Biobank for generously sharing their data. We also wish to thank Prof. Gianpiero Cavalleri and Dr. Vaĺeria Lima Passos for their guidance.

## Supporting information

### Supplementary note 1: Phenotype definitions

#### Type 2 diabetes

Individuals assigned into the positive class were those with any of the following codes in “Diagnoses - main ICD10”, “Diagnoses - secondary ICD10”, or “Diagnoses - ICD10”: “E110”, “E111”, “E112”, “E113”, “E114”, “E115”, “E116”, “E117”, ”E118”, “E119”.

#### Coronary artery disease

Individuals assigned into the positive class were those with any of the following codes in “Diagnoses - main ICD10”, “Diagnoses - secondary ICD10”, or “Diagnoses - ICD10”: “I200”, “I201”, “I208”, “I209”, “I210”, “I211”, “I212”, “I213”, “I214”, “I219”, “I220”, “I221”, “I228”, “I229”, “I235”, “I236”, “I238”, “I240”, “I250”, “I251”, “I252”, “I253”, “I254”, “I255”, “I256”, “I258”, “I259” and “I21.X”

#### Breast cancer

Individuals assigned into the positive class were those with any of the those with self-reported cancer code 1002.

#### Prostate cancer

Individuals assigned into the positive class were those with any of the those with self-reported cancer code 1044.

#### Educational attainment

Codes in UKBB Qualifications Field (6138) were converted to years of education via the following mapping: (1,2,3,4,5,6,-7,-3) *→* (20,13,10,10,19,15,7,NA). From this, a constructed binary phenotype was generated consisting of those with *≤*7 years of education (positive class) and those with *>*7 years of education (negative class). Dichotomizing educational attainment in this manner sacrifices information and interpretability. We employ it only to allow for confounding comparisons across binary disease outcomes. Results for this proxy should not be interpreted as estimates for a true binary educational phenotype.

#### Supplementary note 2: Principal component analysis

All UK-Biobank participants were plotted using their first 16 PCs. Individuals were coloured according to their country or region of origin. Country of birth information were taken from UK Biobank data fields 1647 and 20115. Individuals with missing or unassigned data were excluded. A maximum of 1,000 randomly sampled individuals per country were kept for ease of visualization purposes. For the global plot, all non-European countries were collapsed into four continental bins —Africa, Asia, Americas, Oceania, while the UK and the four core European UN Statistical Division (UNSD) subregions were kept as distinct categories. For the regional plot, only people born in the UK constituent countries and the Republic of Ireland were included.

This analysis of the first 16 principal components derived from UK Biobank SNP data revealed clear evidence of both global and intra-European population structure. Broad ancestry differences were primarily captured by PCs 1, 6, and 11, whilst PCs 4, 7, and 8 reflected finer-scale structure within Europe (Figure S1).

When the analysis was restricted to participants born in the UK and Ireland, additional regional clustering was evident. Although the first four principal components did not appear to pull out specific sub-regions, PCs 5, 9, 11, and 14 showed distinct separation among the constituent countries of the UK and Ireland (Fig. S2).

### Association between PCs and phenotypes

#### GWAS/PRS results with recomputed PCs

The first 16 PCs were re-calculated on the post-QC self-reported white dataset using PLINK2’s approximate PCA method [43, 60]. 126,575 pruned SNPs were used as input for this PCA using a window-size of 50kb, step-size of 5bp and squared correlation threshold of 0.05 [42]. The HLA region was excluded during pruning. A standard GLM-based GWAS and PRS were performed as in the main text using these re-computed 16 PCs as well as age and sex as covariates.

Supplementary note 3: Effects of number of included PCs on the genomic inflation factor ***λ*** and LDSC ratio

Supplementary note 4: Effects of number of included PCs on predicitve performance across all SNP-set sizes

Coronary Artery Disease

Type 2 Diabetes

**Supplementary note 5: Effects of number of included PCs on attenuation across all SNP-set sizes**

**Supplementary note 6: Full GLM and GLMM results**

Breast Cancer

Prostate Cancer

Educational Attainment

---

**Fig S1.**
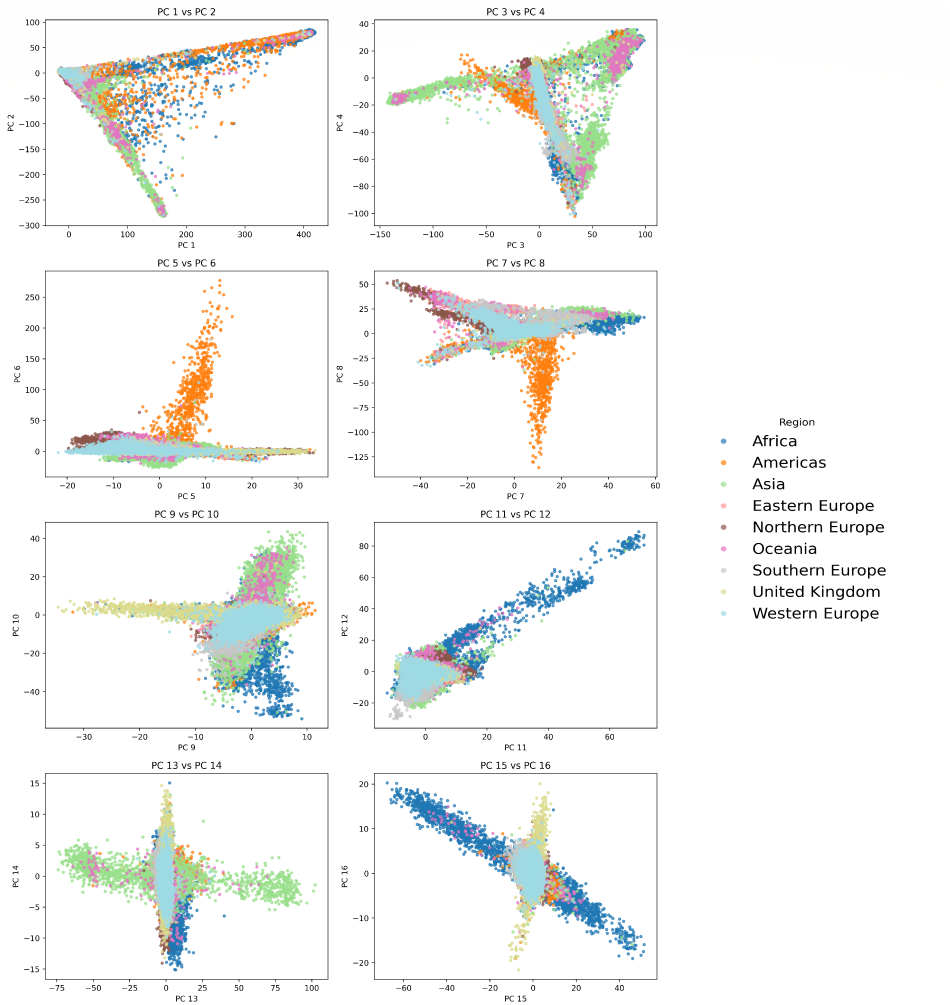
PC Scores 1–16 of UK Biobank Participants PC scores were taken from Data Field 22009 of UK Biobank. A maximum of 1,000 individuals were included per country. Individuals not assigned to any country or population were removed.

**Fig S2.**
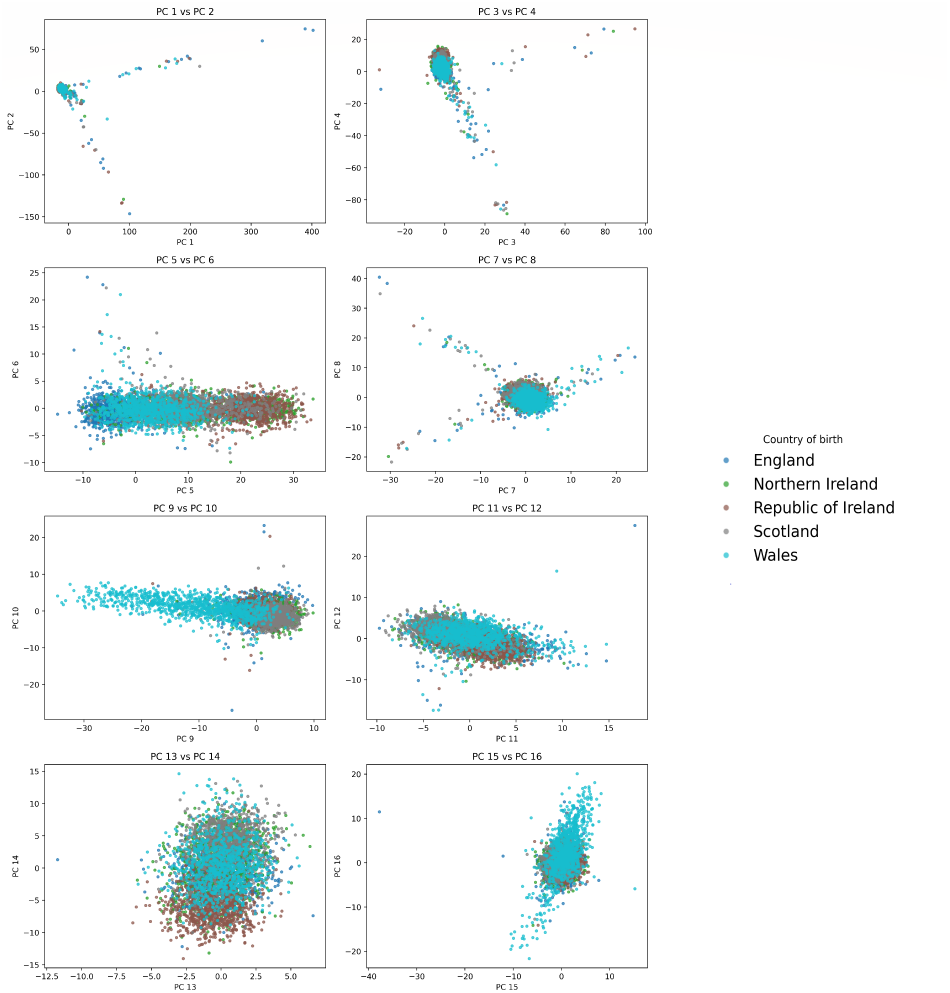
PC Scores 1–16 of UK Biobank Participants (UK and Ireland Only) PC scores were taken from Data Field 22009 of UK Biobank. A maximum of 1,000 individuals were included per country.

**Fig S3.**
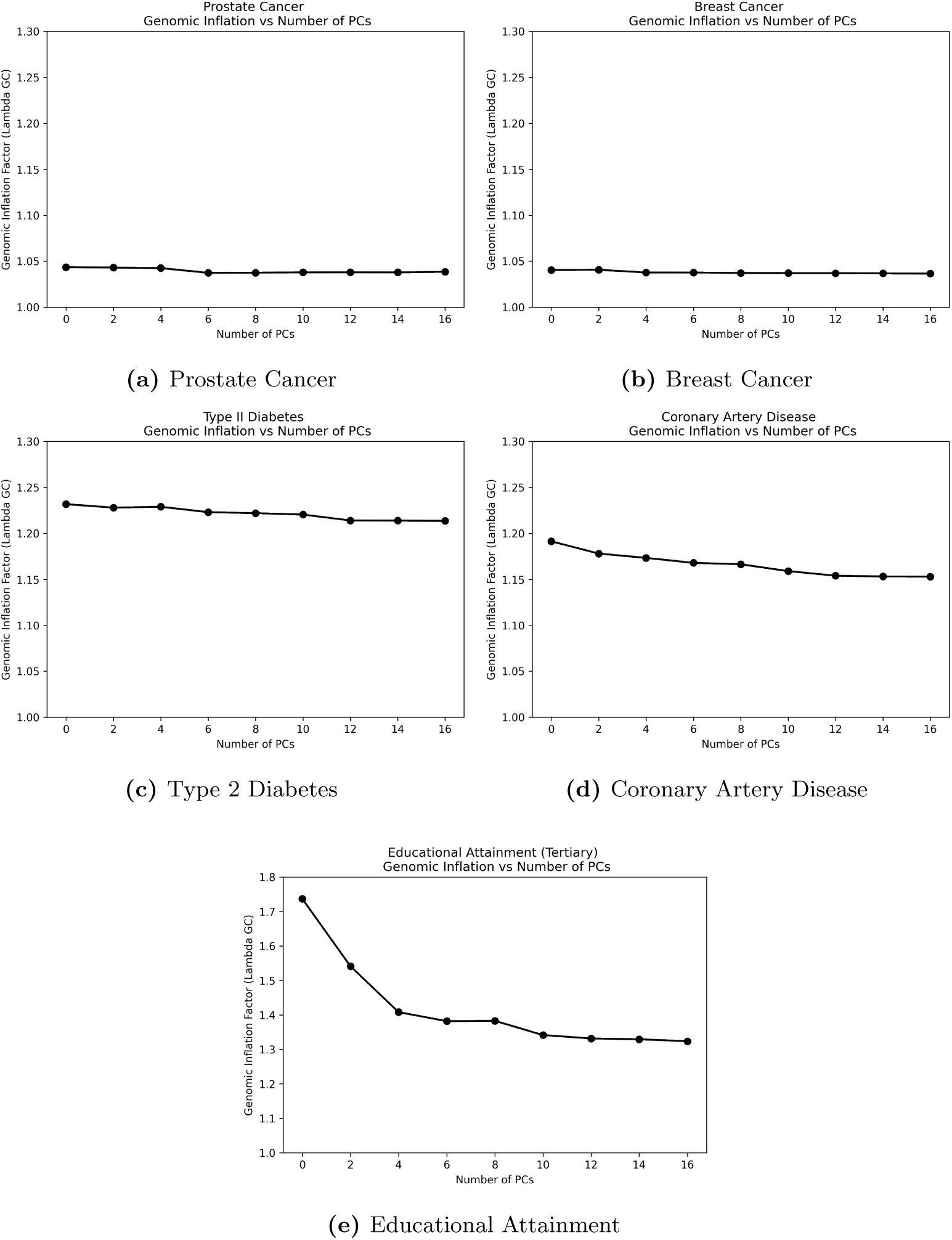
Effects of PCs on the genomic inflation factor *λ*. Summary statistics to calculate *λ* were generated from GLM-based GWAS. Note that the y-axis limit is higher for the educational attainment phenotype due to additional confounding.

**Fig S4.**
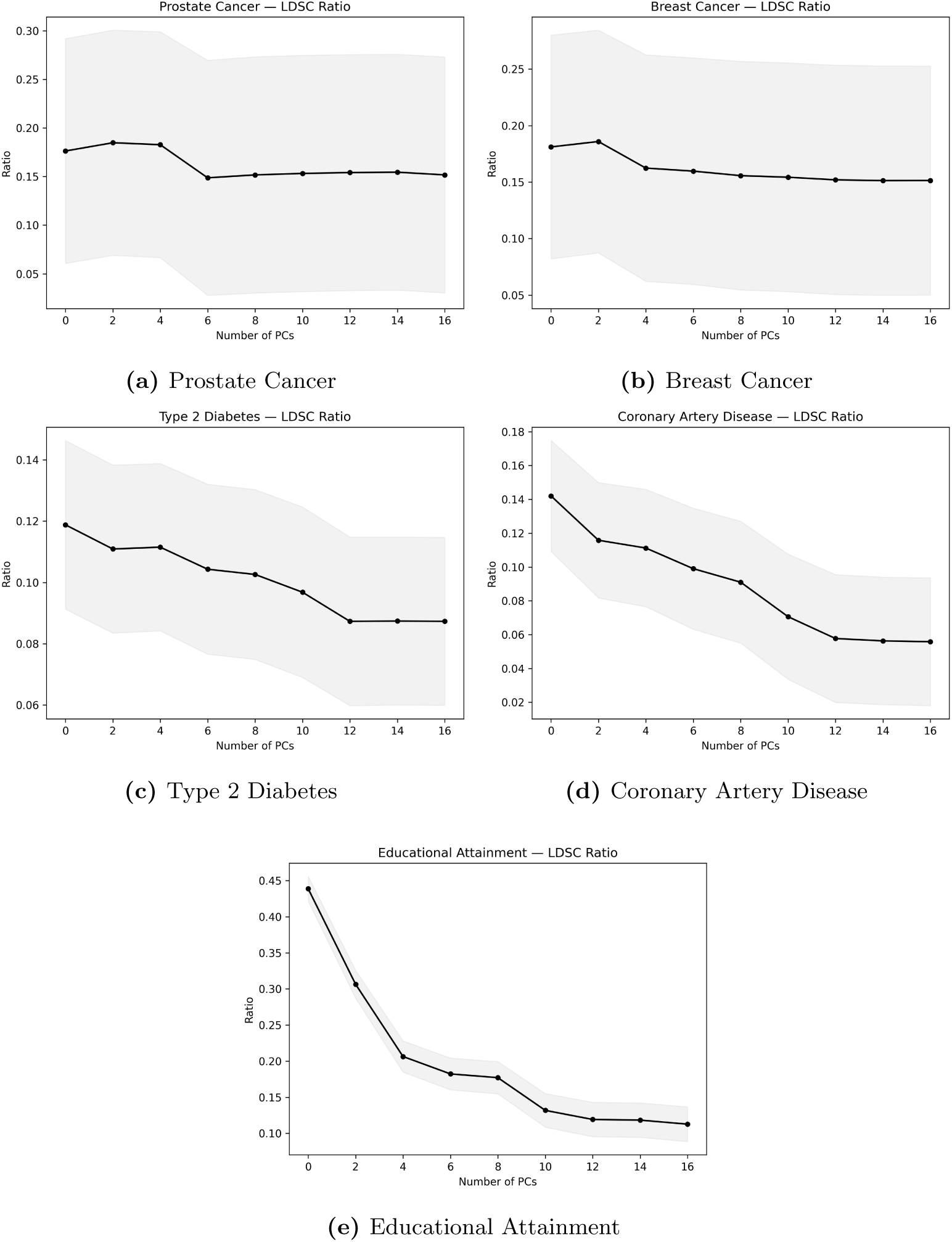
Effect of increasing PC inclusion on the LD-score regression (LDSR) ratio. The ratio is calculated as: (Intercept *−* 1)*/*(mean(*χ*^2^) *−* 1). Summary statistics to perform LDSC were taken from GLM-based GWAS. The standard error is indicated by shading.

**Fig S5.**
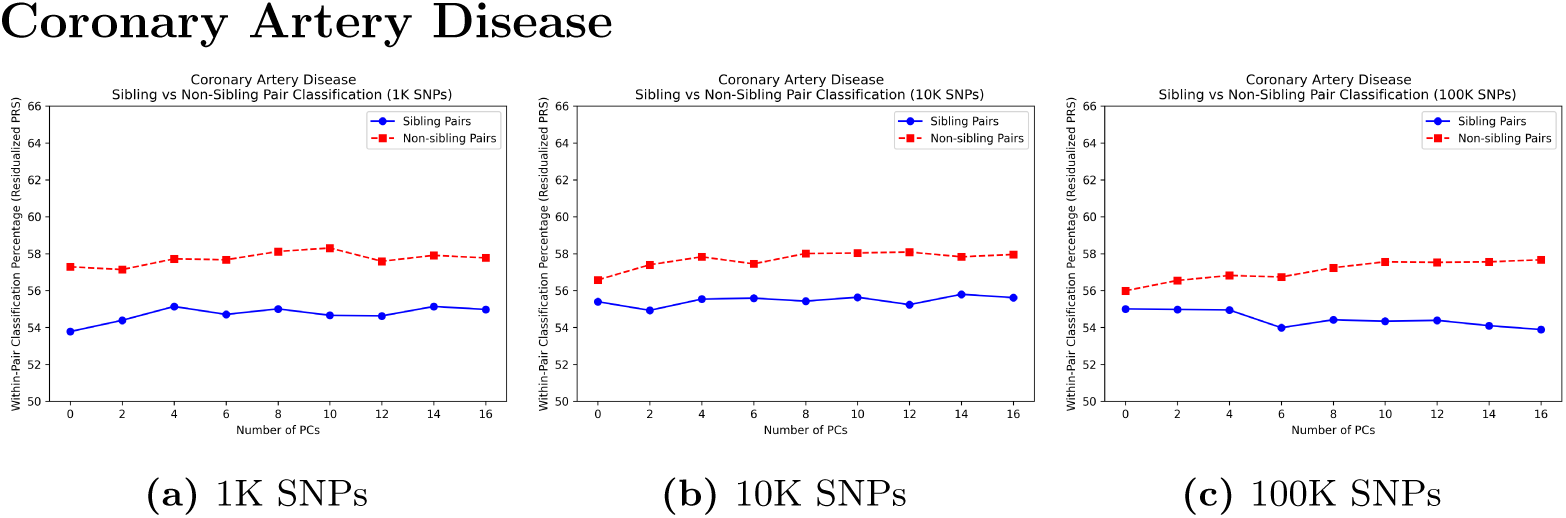
PRS Results for Coronary Artery Disease. PRS were derived from GLM-based GWAS. Predictive performance among sibling and non-sibling pairs with increasing inclusion of PCs is shown.

**Fig S6.**
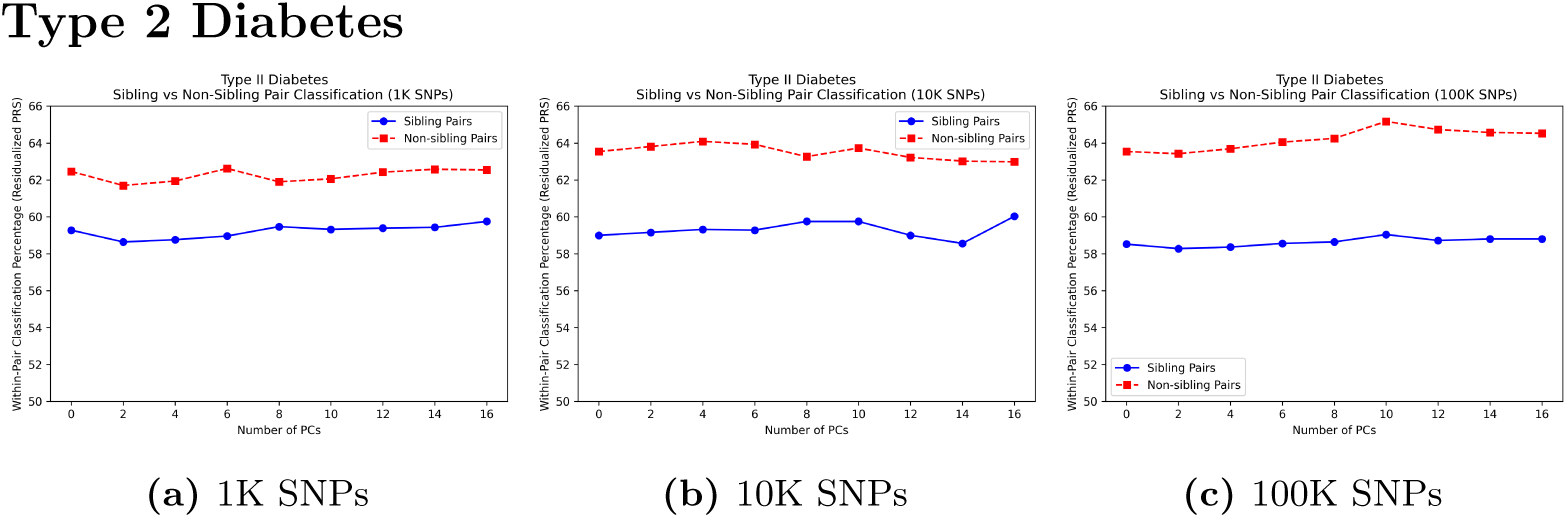
PRS Results for Type 2 Diabetes. PRS were derived from GLM-based GWAS. Predictive performance among sibling and non-sibling pairs with increasing inclusion of PCs is shown

**Fig S7.**
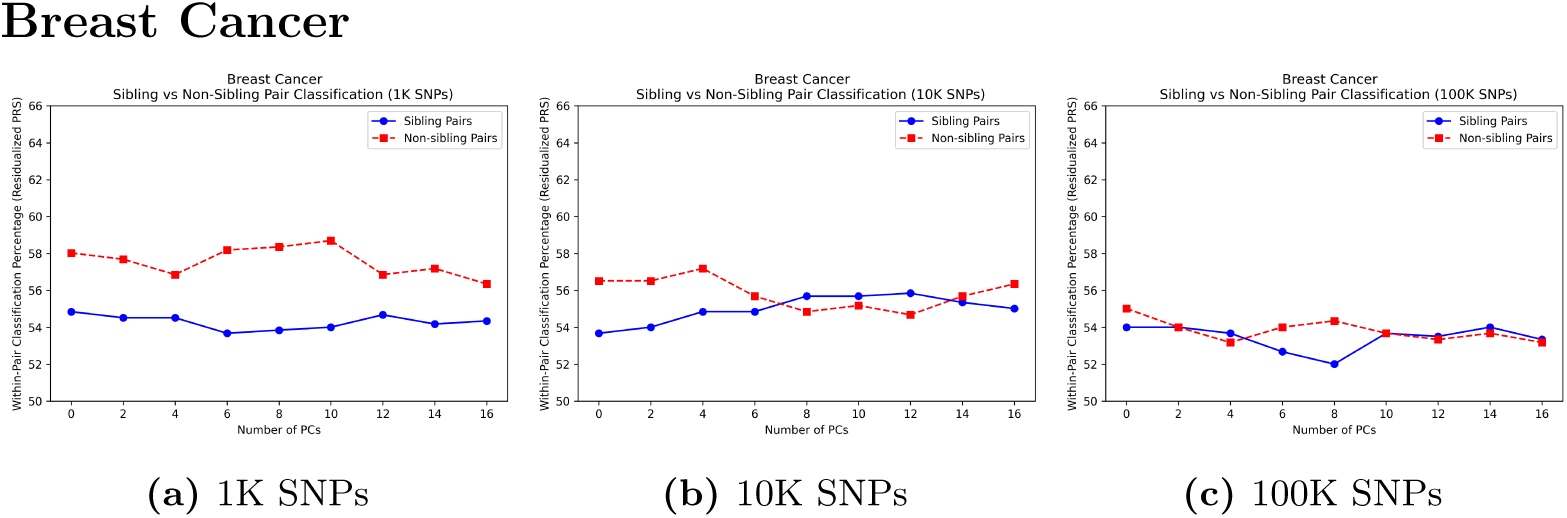
PRS Results for Breast Cancer. PRS were derived from GLM-based GWAS. Predictive performance among sibling and non-sibling pairs with increasing inclusion of PCs is shown

**Fig S8.**
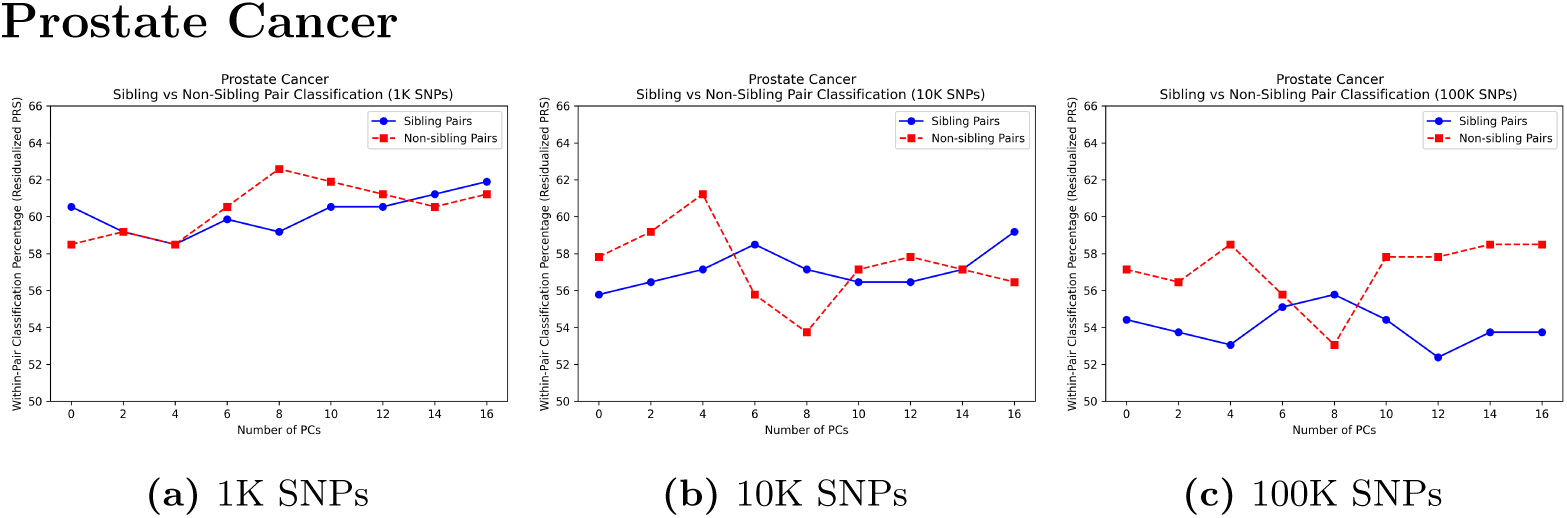
PRS Results for Prostate Cancer. PRS were derived from GLM-based GWAS. Predictive performance among sibling and non-sibling pairs with increasing inclusion of PCs is shown.

**Fig S9.**
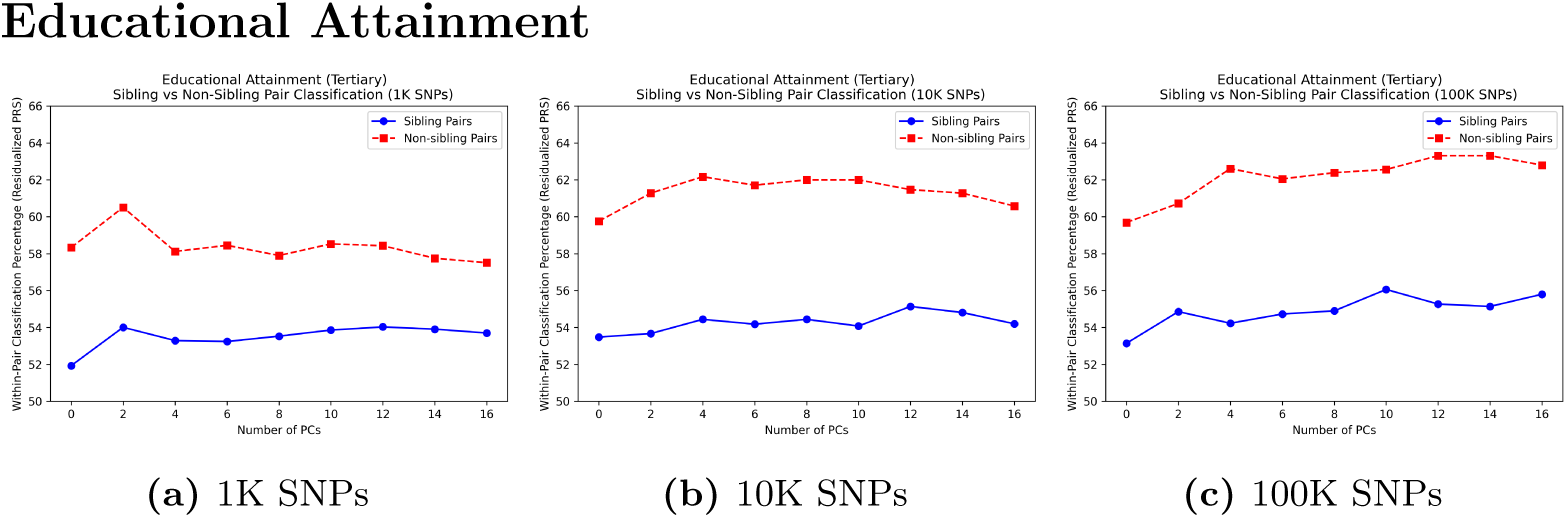
PRS Results for binarized Educational Attainment. PRS were derived from GLM-based GWAS. Predictive performance among sibling and non-sibling pairs with increasing inclusion of PCs is shown.

**Fig S10.**
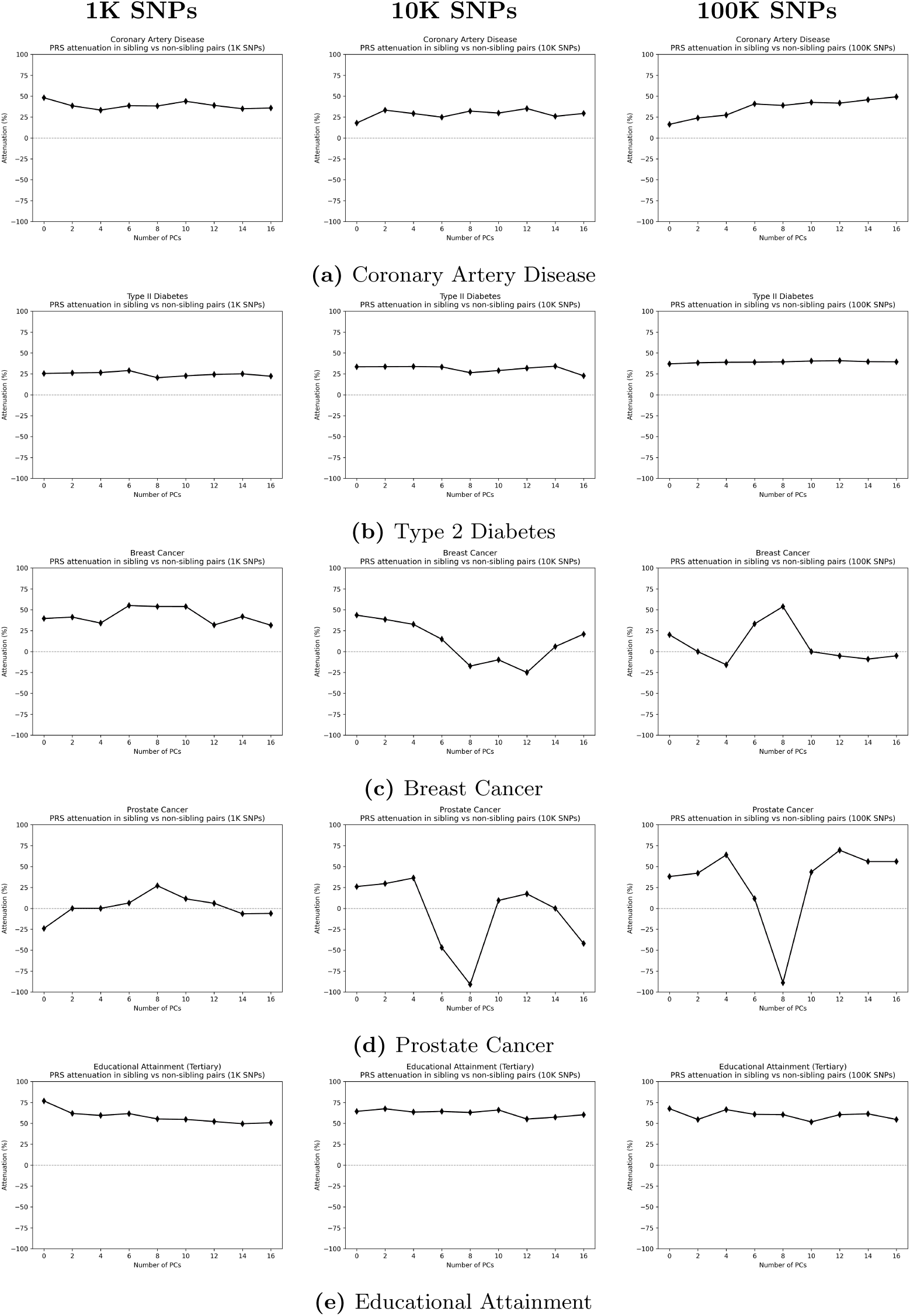
Attenuation in predictive performance with increasing PC inclusion between sibling and non-sibling (population-level) pairs across phenotypes. PRS were derived from GLM-based GWAS.

**Table S1.** Linear regression results between PCs and each phenotype. P-values are shown with significant PCs in bold.

|  | Coronary Artery Disease | Type 2 Diabetes | Prostate Cancer | Breast Cancer | Educational Attainment |
| --- | --- | --- | --- | --- | --- |
| PC1 | 7.7e-01 | **2.0e-03** | 9.6e-01 | 2.1e-01 | **9.7e-05** |
| PC2 | 9.2e-01 | 7.1e-01 | 3.2e-01 | 8.7e-01 | **5.1e-04** |
| PC3 | 1.2e-01 | **5.6e-03** | 1.5e-01 | 3.3e-01 | 8.3e-01 |
| PC4 | **1.4e-04** | **4.4e-02** | 1.6e-01 | 2.7e-01 | **8.6e-50** |
| PC5 | **2.3e-05** | **2.0e-05** | **2.7e-06** | 4.8e-01 | **6.5e-26** |
| PC6 | 1.0e-01 | **4.2e-04** | 6.9e-01 | 1.5e-01 | **3.4e-03** |
| PC7 | 2.8e-01 | **1.7e-02** | 4.4e-01 | 4.9e-01 | **3.0e-48** |
| PC8 | **2.6e-05** | **7.5e-03** | **4.9e-03** | **2.2e-02** | **2.0e-02** |
| PC9 | **4.6e-16** | 5.4e-01 | 9.7e-02 | 9.0e-01 | **3.0e-117** |
| PC10 | **5.8e-03** | 7.3e-02 | 7.5e-01 | 2.3e-01 | **3.3e-03** |
| PC11 | **2.2e-16** | **1.4e-14** | 7.0e-01 | 9.4e-01 | **1.4e-38** |
| PC12 | 1.4e-01 | **3.5e-03** | 9.7e-01 | 5.9e-02 | 7.4e-02 |
| PC13 | 8.2e-02 | 3.1e-01 | 9.9e-01 | 2.0e-01 | 5.4e-01 |
| PC14 | **8.1e-03** | 5.2e-02 | 5.5e-01 | **3.9e-02** | **1.6e-03** |
| PC15 | 9.0e-01 | 3.4e-01 | 2.3e-01 | 3.1e-01 | 3.4e-01 |
| PC16 | **1.9e-02** | **5.5e-04** | 2.1e-01 | 1.3e-01 | **2.4e-31** |

**Table S2.** PRS with recomputed PCs: Classification accuracy (%) in standard GLM-based GWAS-based PRS performance on discordant sibling pairs in the UK Biobank. Age, sex and the first 16 recomputed PCs were included in the GWAS and PRS.

| Phenotype | 1K SNPs | 10K SNPs | 100K SNPs |
| --- | --- | --- | --- |
| Coronary Artery Disease | 56.07 | 55.11 | 56.07 |
| Type 2 Diabetes | 59.24 | 59.75 | 58.28 |
| Breast Cancer | 53.51 | 54.68 | 54.18 |
| Prostate Cancer | 56.46 | 61.90 | 53.74 |

**Table S3.** PRS with recomputed PCs: Classification accuracy (%) in standard GLM-based GWAS-based PRS performance on non-sibling discordant pairs in the UK Biobank. Age, sex and the first 16 recomputed PCs were included in the GWAS and PRS.

| Phenotype | 1K SNPs | 10K SNPs | 100K SNPs |
| --- | --- | --- | --- |
| Coronary Artery Disease | 57.45 | 58.39 | 57.29 |
| Type 2 Diabetes | 62.30 | 63.54 | 64.25 |
| Breast Cancer | 58.36 | 55.69 | 54.35 |
| Prostate Cancer | 61.22 | 56.46 | 61.22 |

**Table S4.** Genomic inflation factor (*λ*) of GLM- and GLMM-based GWAS.

| Phenotype | GLM | GLM+16PC | GLMM+16PC |
| --- | --- | --- | --- |
| Coronary Artery Disease | 1.191 | 1.153 | 1.152 |
| Type 2 Diabetes | 1.232 | 1.214 | 1.211 |
| Breast Cancer | 1.040 | 1.037 | 1.037 |
| Prostate Cancer | 1.043 | 1.039 | 1.036 |
| Educational Attainment | 1.737 | 1.324 | 1.317 |

**Table S5.** Classification accuracy for standard GWAS-based PRS performance on sibling and non-sibling (population-level) discordant pairs in the UK Biobank. The full 16 PCs were included in both the GLM-based GWAS and PRS.

| Phenotype | SNP-set size | Classification accuracy (%) |  |
| --- | --- | --- | --- |
|  |  | Non-sibling pairs | Sibling pairs |
| Coronary Artery Disease | 1K | 57.77 | 54.98 |
|  | 10K | 57.96 | 55.62 |
|  | 100K | 57.67 | 53.89 |
| Type 2 Diabetes | 1K | 62.54 | 59.75 |
|  | 10K | 62.98 | 60.03 |
|  | 100K | 64.53 | 58.80 |
| Breast Cancer | 1K | 56.35 | 54.35 |
|  | 10K | 56.35 | 55.02 |
|  | 100K | 53.18 | 53.34 |
| Prostate Cancer | 1K | 61.22 | 61.90 |
|  | 10K | 56.46 | 59.18 |
|  | 100K | 58.50 | 53.74 |
| Educational Attainment | 1K | 57.51 | 53.70 |
|  | 10K | 60.58 | 54.20 |
|  | 100K | 62.80 | 55.80 |

**Table S6.** Classification accuracy for GLMM-based GWAS PRS performance on sibling and non-sibling (population-level) discordant pairs in the UK Biobank. The full 16 PCs were included in both the GLMM-GWAS and PRS.

| Phenotype | SNP-set size | Classification accuracy (%) |  | Attenuation (%) |
| --- | --- | --- | --- | --- |
|  |  | *Non-sibling pairs* | *Sibling pairs* |  |
| Coronary Artery Disease | 1K | 57.72 | 54.98 | 35.5 |
|  | 10K | 57.77 | 55.62 | 27.7 |
|  | 100K | 57.69 | 53.94 | 48.8 |
| Type 2 Diabetes | 1K | 62.70 | 59.67 | 23.9 |
|  | 10K | 63.02 | 59.83 | 24.5 |
|  | 100K | 64.49 | 59.00 | 37.9 |
| Breast Cancer | 1K | 55.69 | 54.85 | 14.8 |
|  | 10K | 55.02 | 53.68 | 26.7 |
|  | 100K | 55.85 | 54.52 | 6.8 |
| Prostate Cancer | 1K | 59.18 | 61.22 | -22.2 |
|  | 10K | 53.74 | 61.22 | -200.0 |
|  | 100K | 58.50 | 56.46 | -24.0 |
| Educational Attainment | 1K | 57.51 | 53.62 | 51.8 |
|  | 10K | 60.72 | 54.18 | 61.0 |
|  | 100K | 62.87 | 55.58 | 56.6 |

